# Oro-faecal transmission of SARS-CoV-2: A systematic review of studies employing viral culture from gastrointestinal and other potential sources

**DOI:** 10.1101/2024.02.29.24303532

**Authors:** Sara Gandini, John Conly, Elizabeth A. Spencer, David Evans, Elena C Rosca, Jon Brassey, Susanna Maltoni, Igho Onakpoya, Annette Plüddemann, Tom Jefferson, Carl Heneghan

## Abstract

**Background:** The extent to which the oro-faecal route contributes to the transmission of SARS-CoV-2 is not established.

**Methods:** We systematically reviewed the evidence on the presence of infectious SARS-CoV-2 in faeces and other gastrointestinal sources by examining studies that used viral culture to investigate the presence of replication-competent virus in these samples. We conducted searches in the WHO Covid-19 Database, LitCovid, medRxiv, and Google Scholar for SARS-CoV-2 using keywords and associated synonyms, with a search date up to 28th of November 2023.

**Results:** We included 13 studies involving 328 COVID-19 subjects - providing 314 faecal or rectal swab SARS-CoV2 positive samples tested also with viral culture. The methods used for viral culture across the studies were heterogeneous. Three studies (2 cohorts and 1 case-series) reported observing replication-competent SARS-CoV-2 confirmed by quantitative RT-PCR (qPCR) and whole genome sequencing, and qPCR including appropriate cycle threshold changes. Overall, six (1.9%) of 314 faecal samples subjected to cell culture showed replication-competent virus. One study found replication competent samples from one immunocompromised patient. No studies were identified demonstrating direct evidence of oro-faecal transmission to humans.

**Conclusions:** Our review found a relatively low frequency of replication-competent SARS-CoV-2 in faecal and other gastrointestinal sources. Although it is biologically plausible, more research is needed, using standardized cell culture methods, control groups, adequate follow-up and robust epidemiologic methods, including whether secondary infections occurred, to determine the role of the oro-faecal route in the transmission of SARS-CoV-2.

## Introduction

Human coronaviruses have been shown to affect the human gastrointestinal (GI) tract: MERS-CoV (Middle East respiratory syndrome coronavirus infection) was demonstrated to infect the gastrointestinal tract, with intestinal epithelial cells supporting replication-competent virus [1], and SARS-CoV-1 was associated with gastrointestinal symptoms and prolonged RNA shedding in faeces as by demonstrated by the presence of replication-competent virus in cell cultures of faecal samples from affected patients [2].

It was shown early in 2020 that the SARS-CoV-2 virus uses the angiotensin-converting enzyme 2 (ACE2) receptor as a cell entry receptor to enter ACE2-expressing cells [3]. Since the ACE2 receptor is abundant not only in lung epithelial cells but is also highly expressed on the luminal surface of intestinal epithelial cells [4], a potential route for infection via the gastrointestinal system is considered biologically plausible. Studies have demonstrated that human gastrointestinal tract cell lines with a brush border and colon-derived cell lines plus colonic organoids have robust viral growth after inoculation of SARS-CoV-2 and allow for persistent infection within these cell lines [5–7]. There have also been animal models, including non-human primates which have been inoculated by intragastric intubation, bypassing the respiratory tract, demonstrating unequivocal evidence for direct invasion of the GI tract by SARS-CoV-2 with concomitant GI and lung pathology [8–10]. Given the findings that SARS-CoV-2 is able to replicate within human gastrointestinal tract cell lines and remain infectious on excretion and the establishment of invasive infection in multiple animal models[8]–[10], it was reasonable to consider whether infectious SARS-CoV-2 could be found in faecal specimens or other sources representing potential oro-faecal sources, creating an orofecal transmission risk (also commonly referred to as faecal-oral) [11], which would be important for the application of appropriate infection control measures.

Involvement of the GI tract was one of the reasons for considering that the real rate of paediatric cases may have been higher than that officially reported, as children may present with only GI symptoms and signs SARS-CoV-2 RNA has been identified in anal/rectal swabs and in stool specimens of COVID-19 patients even after upper respiratory tract virus clearance [12]. In contrast to what has been observed in adult patients, higher proportions of fever, vomiting, and diarrhoea have been recorded on admission in paediatric cases [13]. GI localization in children may represent an alternative site of viral shedding and transmission. For these reasons, sanitation measures in schools included interventions directed against oro-faeacal transmission routes [14].

Previously, we systematically reviewed studies reporting on the possibility of transmission via the oro-faecal route and found several studies reporting that SARS-CoV-2 RNA can be shed from the GI tract [15]. However, the shedding of viral RNA does not necessarily equate to viral replication in the GI tract. Since that report, it has become more apparent that understanding the transmission of SARS-CoV-2 depends on studies using high-quality, replicable methods to assess the potential infectivity of samples, along with rigorous epidemiological data examining exposures and outcomes [16]. We therefore set out to identify, appraise, and summarise the evidence on the presence of replication-competent SARS-CoV-2 in human faecal and other source specimens that could represent faecal-oral transmission pathways, and if cases of human oro-faecal transmission of SARS-CoV-2 have been convincingly demonstrated.

## Materials and Methods

We performed this systematic review following our published protocol[17]. In brief, we searched for studies reporting data from participants with COVID-19 (with or without control groups) from whom biological samples were obtained. We verified whether there was a laboratory confirmation of SARS-CoV-2, and corresponding clinical data, including symptomatology, disease course, treatments, and comorbidities. We focused on studies reporting results of viral culture from human faecal and other gastrointestinal (GI) samples, coupled with reverse-transcription polymerase chain reaction (RT-PCR) testing, to investigate what is known about the presence of replication-competent virus within faecal samples from individuals with COVID-19 and the demonstration of any onward orofaecal transmission of SARS-CoV-2. To be included studies must have complied with all the following criteria: included viral culture of faecal/GI samples from SARS-CoV-2 infected persons, and assessed the cytopathic effect and verification techniques of the isolated virus to ensure the cultured virus was SARS-CoV-2. Studies with data only from RT-PCR testing of faecal samples from SARS-CoV-2 infected persons and/or reporting solely predictive modelling were excluded. In order to help reduce bias due to selective reporting, case studies of one or two cases were excluded as well.

We assessed the risk of bias within five domains, modified and extended from the Quality Assessment of Diagnostic Accuracy Studies (QUADAS-2) criteria [18]. Further details regarding methods are reported in the supplementary materials (Appendix 1).

## Results

Figure 1 shows the flow of studies through the screening and inclusion/exclusion processes. The literature searches identified 1496 studies, and 14 were found from other sources, even if some were duplications. After screening abstracts, 53 independent studies were found eligible. We excluded 40 studies for various reasons (Appendix 2. Studies excluded on full-text screening). Finally, we included 13 studies with a total of 1625 participants [19–31].

**Figure 1.**
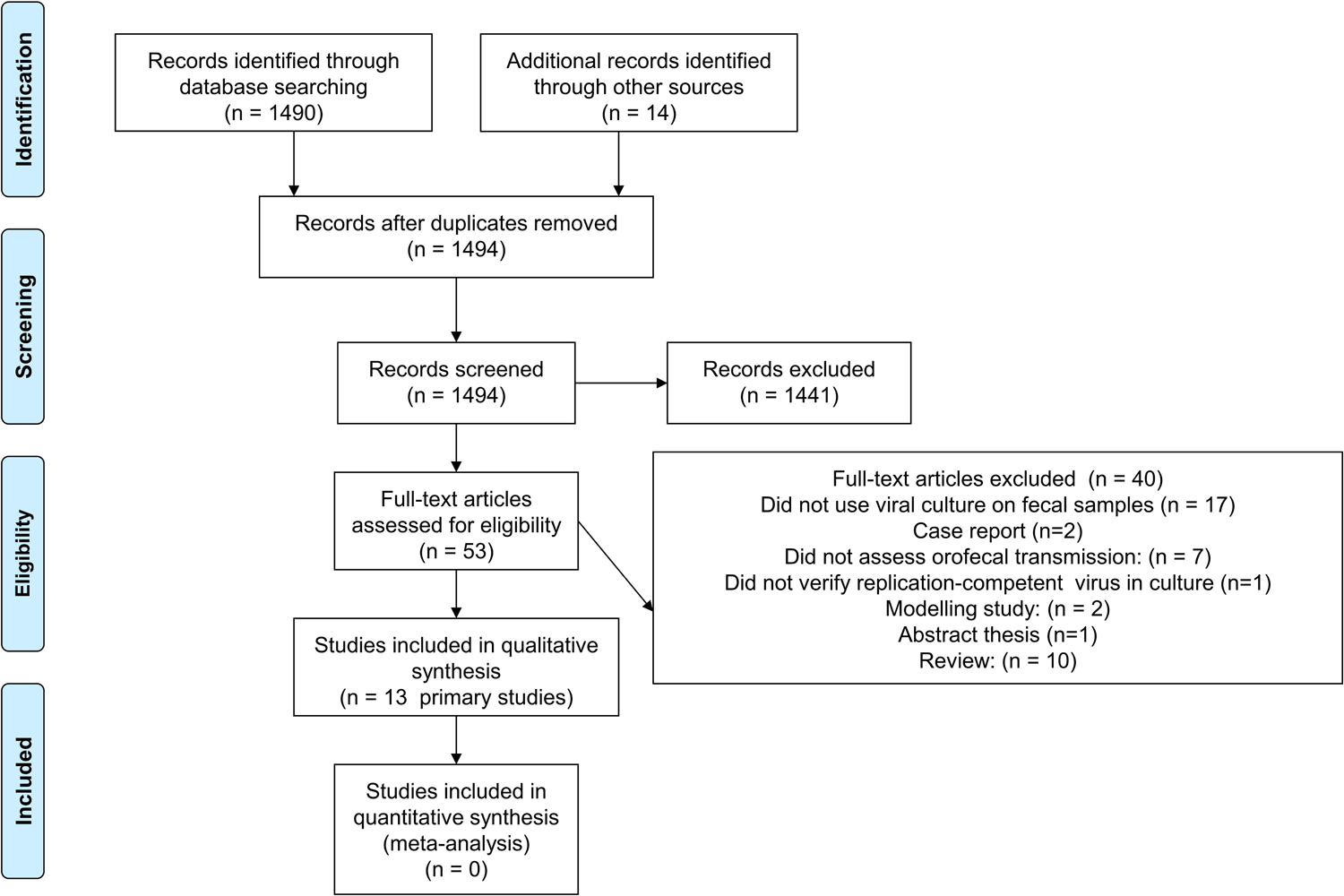
Flow diagram showing the process for inclusion/exclusion of studies.

Table 1 shows the main characteristics of the included studies and viral culture information on SARS-CoV-2-positive subjects. More detailed information is presented in Tables S1. Eight of the studies were cohort, while the remaining five were case series. The 13 studies involved mainly hospitalised patients but also some persons from ambulatory or home settings. Seven studies were conducted in Europe, four in Asia, and two in South America. Study duration ranged from two weeks to 15 months.

**Table 1.** Descriptive features of included studies and results from samples for cell culture.

| First Author, PY | Study design | Calendar date/period | Country | No. individuals with samples used for culture* | No. samples tested for viral culture | No. with Ct value < 30 at baseline | No. positive samples from cell culture† |
| --- | --- | --- | --- | --- | --- | --- | --- |
| Akiyama 2022 | Cohort study | January to March 2020 | Japan | 4 | 8 | NR | 0 |
| Albert 2021 | Cohort study | June 2020 to December 2020 | Spain | 7 | 6 | 0 | 0 |
| Cerrada-Romero 2022 | Cohort study | March 2020 to February 2021 | Spain | 27 | 31 | NR* | 0 |
| Dergham 2021 | Cohort study | March 2020 to April 2020 | France | 46 | 106 | 0† | 2† |
| Fumian 2021 | Case series | January to July 2020 | Brazil | 4 | 4 | 1 | 0 |
| Jeong 2020 | Cohort study | February 2020 to March 2020 | S Korea | 3 | 3 | NR | 0** |
| Joshi, 2022 | Case series | June to August 2021 | India | 10 | 10 | 10 | 0 |
| Lavania 2022 | Cohort study | May 2020 to August 2021 | India | 55 | 55 | NR € | 0 |
| Nogueira, 2022 | Case series | December 2020 and June 2021 | Austria | 7 | 20 | 13 | 0 |
| Pedersen 2022 | Cohort study | October 2020 to March 2021 | Denmark | 28 | 33 | 3 | 0 |
| Ribeiro, 2022 | Cohort study | May to October 2020 | Brazil | 130 | 14 | 14 | 1 |
| Wölfel 2020 | Case series | January 2020 | Germany | 4 | 13 | NR | 0 |
| Yao 2020 | Case series | 22 January to 4 February 2020 | China | 3 | 11 | 3 | 3 |

† See supplementary materials for verification methods used. \*faecal samples, rectal swabs, wastewater or other GI related samples. ¥ rectal swabs. PY: publication year. NR: Not Reported. € A previous protocol stated only done if < 30. For Albert et al the number of samples used for viral culture was estimated from the manuscript but was not clear.

A Japanese case series of 13 COVID-19 patients diagnosed between January and March 2020 reported data of viral load from faeces by days and symptoms [19]. One study reported on a series of 6 patients from a study investigating samples from patients in Spanish hospitals and local wastewater samples [20]. One was a prospective multicenter Spanish cohort, which analysed 31 samples obtained from 27 patients within two weeks of admission with COVID-19 symptoms between March 2020 and February 2021 [24]. One was a case series from a hospital in France reporting on 46 patients admitted with COVID-19 between March and April 2020 from whom 106 stool samples were taken as part of standard care; viral culture was reported for just one patient with kidney transplantation under immunosuppressive therapy who was admitted for severe diarrhoea [25]. One study from a hospital in Brazil presented a retrospective case series using stored 4 samples collected from 4 patients between January and July 2020 for ongoing surveillance of acute gastroenteric disease[26]. A small Korean study presented data collected from five COVID-19 patients and performed a quantitative polymerase chain reaction (qPCR) to assess viral load[27]. Specimens positive with qPCR were subjected to virus isolation in Vero cells and they reported log copy numbers for faecal samples from three patients. Joshi presented a multicenter case-series from hospitals in India using stored samples collected between May 2020 and August 2021[28]. A further study from India described a multicenter cohort study involving 55 patients (55 samples) attending hospitals who tested positive for SARS-CoV-2 between May 2020 and August 2021[29]. A study from three clinical centres in Austria reported data on 206 hospitalised children diagnosed between December 2020 and June 2021[30]. A study from Denmark reported data on 28 patients diagnosed between October 23, 2020, and March 17, 2021[31]. Another cohort study [21] described data from a slum in Rio de Janeiro, Brazil, collected from May to October 2020, regarding the SARS-CoV-2 test results of 333 rectal swabs from 130 persons, some of whom were symptomatic patients and others were their household contacts. A cluster of epidemiologically linked 4 cases (corresponding to 13 samples) managed in hospitals in Germany early in the pandemic was reported [22]. Finally, one Chinese study analysed data from stool samples of 3 of 11 patients diagnosed between January 22 to February 4, 2020 [23].

Across these 13 studies, there were an estimated 324 total samples from 328 hospitalised patients, households and other settings (exact numbers were not always clearly reported) analysed with viral culture. Among these, three studies reported finding evidence of an isolated virus (Table 1); which were confirmed positive by qPCR and/or genome sequencing of SARS-CoV-2 in the culture supernatant. One of these three studies was a cohort study, in which the positive samples identified by viral culture were obtained from an immunocompromised patient. The samples were obtained at variable time points during the course of illness, and only nine studies reported Ct values from the stool specimens at the time of initial attempts at culture. Of the studies that reported Ct values from the faecal and other samples, all the 6 samples found to have presumptive evidence of culturable virus had Ct values reported as ≤ 30, which is in the range where a high likelihood of culturable virus may be found [22], [32], [33]. Considering only the cohort studies, there was a 1.2% (3/256 samples) positive frequency for those specimens.

Overall, we identified a low-moderate risk of bias across the included studies in four domains but a high risk of bias in one domain, with evident deficits in reporting viral cell culture methods.

Table S2 shows the risk of bias assessment results. The risk of bias was generally considered low for the criteria for the domains of diagnosing a case clearly and appropriately, reporting of patient/ population characteristics adequacy and reporting of methods used to obtain RT-PCR results replicable and appropriate. Reporting of patient characteristics was absent or unclear in only one (8%) studies and methods for RT-PCR were unclear in four (31%) studies. However, the risk of bias was considered high for clear reporting of viral culture methods, where it was only well reported in five studies. Analysis and appropriate reporting of the results was considered to have low-moderate risk of bias with unclear reporting in two studies.

## Discussion

In this systematic review, we included 13 cohort and case-series studies that subjected faecal, rectal swabs or other GI samples (from hospitalised patients and persons in the community with confirmed COVID-19) to viral culture. Among a total of 314 samples subjected to cell culture which were found to be RT-PCR + for SARS-CoV-2, six (1.9%) were reported as positive from cell culture. The sampling methodology was poorly described in some studies and appeared to be more of a convenience sampling as opposed to a rigorous hypothesis driven type of research protocol, with some exceptions. Sampling stool at late stages of illness with no attention paid to cycle threshold values or quantitative viral load would lead to a large number of samples that would have a high likelihood of being negative for culture given our knowledge about the natural history of COVID-19 [22, 32, 33].

Schedules of sample collection and information on symptoms across the disease course were incompletely reported. Coupled with the limited number of data points available, it was not possible to assess the relationship between the time of symptom onset, faecal sample collection, and results of viral culture. The timing of sample collection is likely to be important[34], and most of the studies reported herein are too few, varied, and unclear in their descriptions to be able to determine whether the included participants were indeed within a potential infectious window. However, the high Cts of many of the faecal and respiratory samples reported suggest that most of the included subjects were unlikely to still be infectious at the time of faecal sample collection. It is also possible that SARS-CoV-2 typically has only a brief survival time with transit through the GI tract or after exiting it; this is as yet unknown.

Nonetheless, in those samples where attention was paid to the timing of faecal sample collection with respect to the course of the illness and/or with low Ct values, higher frequencies of virus isolation were reported. It is possible these current studies underrepresent the presence of culturable virus in human stool specimens due to poor sampling strategies.

The likelihood of transmission via the oro-faecal route is definitely biologically plausible from evidence on other human and animal coronaviruses, including MERS-CoV and SARS-CoV-1 [1,2, 36] long with studies early in the pandemic reporting findings that suggested entry of SARS-CoV-2 into gastrointestinal cells and possible replication with the GI tract, along with many experimental animal model studies [8–[0].

In addition, Qian et al. [37] reported a study using electron microscopy to examine tissue obtained from the rectal mucosa of a patient undergoing rectal surgery who subsequently developed COVID-19 symptoms. The authors reported observing virions matching SARS-CoV-2 morphology in sections of the obtained tissue sample. As described above, however, viral morphology alone is not a sufficient indicator of identity [38], and in itself does not show that replication-competent virus is present; this observation could not be regarded as sufficient to indicate a transmission risk from faeces.

In February 2020 Zhang and colleagues [13] reported testing stool samples by cell culture in Vero cells. The authors reported on the finding of successful cell culture in one sample taken from a laboratory-confirmed COVID-19 case with severe pneumonia, 15 days after the onset of symptoms. They reported that viral particles with morphology typical of SARS-CoV-2 were observed under electron microscopy; no further details were reported.

To assess a chain of transmission for SARS-CoV-2, it is necessary to have reliable epidemiology, adequate reporting of symptoms and signs, sufficient follow-up, and evidence from human samples of the presence of a replication-competent virus[39]. We previously defined viral culture as encompassing several methods that can uniquely identify the replicating agent as SARS-CoV-2 [40]. Most commonly, this would be a plaque assay combined with an RT-PCR diagnosis or immunological staining, or gene sequencing of viral RNA.

Numerous studies have identified SARS-CoV-2 RNA in faecal samples and wastewater[5], suggesting the possibility of infectious virus remaining in faeces. Although useful for surveillance purposes, RT-PCR positivity in wastewater is of little significance in understanding transmission without viral load data and cultivability[9]. To the best of our knowledge, no review of the results of viral culture on faecal samples exists.

### Strengths and limitations

We attempted to collate and synthesise all the relevant data from studies using viral culture of faecal samples to investigate the presence of replication-competent virus, which would suggest the potential for oro-faecal transmission of SARS-CoV-2. We used robust methods to search for the relevant studies, and we accounted for the reporting quality of included studies. However, studies investigating the presence of replication-competent virus within faecal samples have been few and have varied in design and methods. Methodologies have generally been lacking in scientific rigour. Comparison between studies is hindered by the lack of standard approaches for sampling schedules, RT-PCR cut-offs, and viral culture methods. In addition, there are acknowledged methodological difficulties in culturing virus from faecal samples due to the presence of heavy bacterial contamination and the presence of inhibitors in stool or direct cytotoxicity of stool specimens [41].

Viral culture has been reported infrequently in respiratory samples with cycle thresholds above 25 [34], and it is likely to assume similar results within faecal samples with high Ct values. Selecting samples with a RT-PCR cycle count of higher than 25 greatly reduces the likelihood of being able to culture SARS-CoV-2 [32, 39, 42]. In the studies in this review, in general, samples with relatively high RT-PCR cycle counts were subjected to viral culture testing.

Standardised, appropriate methods for viral culture studies are essential to generate reliable evidence that can be compared between studies and allow the combining of results. A weakness in our review was the reporting of rigorous virologic methods and use of controls. Only two studies [25, 29] reported using negative/uninfected controls for the cell culture experiments; in no study were methods to reduce contamination described; and no study demonstrated an increasing viral load within cell culture supernatants related to a clear timeline and course of the disease.

### Implications for research and policy

It is difficult to draw firm conclusions about the likelihood of infectiousness of faecal samples and the associated potential transmission via the oro-faecal route, given the limitations of the available data. Although our review suggests the presence of infectious virus appears to be of low frequency (1.9%) from these samples, it is important to recognise that not accounting for settings whereby Ct values are above a threshold allowing for a high likelihood of negative cultures and with testing in a non-standardized manner, the results are likely an underestimate. In addition, the study by Ribeiro et al [21] found the presence of replicating SARS-CoV-2 in 19.4% (6/31) of samples tested, measured by the detection of sgN mRNA, suggesting the degree of infectious virus may be higher than that found by cell culture and this approach deserves further exploration.

Our findings are consistent with the findings in a recent systematic review whereby 8.3% of fomite samples which were RT-PCR + were found to have infectious SARS-CoV-2 [41]. They also reported that the highest frequency of detection was within seven days of symptom onset and significantly associated with a Ct< 30. Placing our findings in context, when extrapolated to millions of COVID-19 cases globally, the real potential for an orofecal route of transmission in high risk settings deserves more attention. However, it remains biologically plausible and this route of potential transmission needs further exploration. Given the stability and survivability of SARS-CoV-2 from environmental sources, taken together with our findings demonstrating the presence of infectious virus in 1.9% of faecal and/or rectal samples from humans, a network meta-analysis finding that meal-gathering by individuals had one of the highest risks associated with transmission of SARS-CoV-2 and the recent findings of infectious virus on several types of frozen foods, deli foods, meat, seafood and fresh produce, and ice cream for prolonged periods of time from days to weeks, all add impetus to further explore this route of transmission [25, 43, 44].

It is important to address the possibility of oro-faecal transmission to develop effective infection control measures, particularly in high-risk settings such as hospitals, residential care settings where facilities may be shared and households care workers where a number of at-risk individuals may occur. Evidence from studies using rigorously performed viral culture is needed to investigate the presence of replication-competent virus and relate this to cycle thresholds or other surrogates of infectivity to assess the likelihood of transmission of SARS-CoV-2 via the oro-faecal route.

## Conclusions

This review concludes there is evidence of infectious SARS-CoV-2 in faeces and/or rectal swabs and other GI specimens in the settings described. Our review highlights the need for further high-quality research, ideally from prospective cohort studies, using appropriate timing of specimen capture and using standardised high-quality methods for viral culture with appropriate negative controls, coupled with robust evidence on possible transmission events to ascertain the likelihood of transmission of SARS-CoV-2 via the oro-faecal route. Animal models and human challenge studies are both viable options to help address this research gap.

## Supporting information

Appendix 1. Literature search strategy. App 2. List of excluded studies, with reasons. App 3. Additional information on the methods and results

Table S1 Main findings of the included studies.

## Supplementary Materials

Table S1 Main findings of the included studies.

Table S2 Risk of bias assessment results.

Appendix 1. Literature search strategy.

Appendix 2. List of excluded studies, with reasons.

Appendix 3. Additional information on the methods of the included studies and risk of bias assessment

## Institutional Review Board Statement

No ethics committee approval was necessary for this study because all data used were previously publicly available.

## Data Availability

All data produced in the present work are contained in the manuscript

## References

[1] J. Zhou et al., “Human intestinal tract serves as an alternative infection route for Middle East respiratory syndrome coronavirus,” Sci. Adv., vol. 3, no. 11, 2017, doi: 10.1126/SCIADV.AAO4966.

[2] W. K. Leung et al., “Enteric involvement of severe acute respiratory syndrome - Associated coronavirus infection,” Gastroenterology, vol. 125, no. 4, pp. 1011–1017, 2003, doi: 10.1016/j.gastro.2003.08.001.

[3] P. Zhou et al., “A pneumonia outbreak associated with a new coronavirus of probable bat origin,” Nature, vol. 579, no. 7798, pp. 270–273, 2020, doi: 10.1038/S41586-020-2012-7.

[4] H. Zhang, J. M. Penninger, Y. Li, N. Zhong, and A. S. Slutsky, “Angiotensin-converting enzyme 2 (ACE2) as a SARS-CoV-2 receptor: molecular mechanisms and potential therapeutic target,” Intensive Care Med., vol. 46, no. 4, pp. 586–590, 2020, doi: 10.1007/S00134-020-05985-9.

[5] M. Guo, W. Tao, R. A. Flavell, and S. Zhu, “Potential intestinal infection and faecal-oral transmission of SARS-CoV-2,” Nat. Rev. Gastroenterol. Hepatol., vol. 18, no. 4, pp. 269–283, 2021, doi: 10.1038/S41575-021-00416-6.

[6] S. Lee, G. Y. Yoon, J. Myoung, S. J. Kim, and D. G. Ahn, “Robust and persistent SARS-CoV-2 infection in the human intestinal brush border expressing cells,” Emerg. Microbes Infect., vol. 9, no. 1, pp. 2169–2179, 2020, doi: 10.1080/22221751.2020.1827985.

[7] R. K. Yantiss et al., “Intestinal Abnormalities in Patients With SARS-CoV-2 Infection: Histopathologic Changes Reflect Mechanisms of Disease,” Am. J. Surg. Pathol., vol. 46, no. 1, pp. 89–96, 2022, doi: 10.1097/PAS.0000000000001755.

[8] L. Jiao et al., “The Gastrointestinal Tract Is an Alternative Route for SARS-CoV-2 Infection in a Nonhuman Primate Model,” Gastroenterology, vol. 160, no. 5, pp. 1647–1661, 2021, doi: 10.1053/J.GASTRO.2020.12.001.

[9] A. C. Y. Lee et al., “Oral SARS-CoV-2 Inoculation Establishes Subclinical Respiratory Infection with Virus Shedding in Golden Syrian Hamsters,” Cell reports. Med., vol. 1, no. 7, 2020, doi: 10.1016/J.XCRM.2020.100121.

[10] S. H. Sun et al., “A Mouse Model of SARS-CoV-2 Infection and Pathogenesis,” Cell Host Microbe, vol. 28, no. 1, pp. 124–133.e4, 2020, doi: 10.1016/J.CHOM.2020.05.020.

[11] J. M. van Seventer and N. S. Hochberg, “Principles of Infectious Diseases: Transmission, Diagnosis, Prevention, and Control,” in *International Encyclopedia of Public Health*, Elsevier Inc., 2016, pp. 22–39.

[12] M. G. Puoti, A. Rybak, F. Kiparissi, E. Gaynor, and O. Borrelli, “SARS-CoV-2 and the Gastrointestinal Tract in Children,” Front. Pediatr., vol. 9, 2021, doi: 10.3389/FPED.2021.617980.

[13] C. Zhang et al., “Clinical and epidemiological characteristics of pediatric SARS-CoV-2 infections in China: A multicenter case series,” PLoS Med., vol. 17, no. 6, 2020, doi: 10.1371/JOURNAL.PMED.1003130.

[14] M. Dewan, N. Sharma, P. Panda, and P. Banerjee, “School reopening: Back to classroom. A systematic review of strategies and their implementation during COVID-19 pandemic,” J. Fam. Med. Prim. care, vol. 11, no. 8, p. 4273, 2022, doi: 10.4103/JFMPC.JFMPC_23_22.

[15] E. A. Spencer et al., “SARS-CoV-2 and the role of orofeacal transmission: A systematic review,” F1000Research, vol. 10, 2021, doi: 10.12688/F1000RESEARCH.51592.2/DOI.

[16] T. Jefferson et al., “A Hierarchical Framework for Assessing Transmission Causality of Respiratory Viruses,” Viruses, vol. 14, no. 8, 2022, doi: 10.3390/V14081605.

[17] C. Heneghan et al., “Orofaecal transmission of SARS-CoV-2 and infection with COVID-19: A protocol for a systematic review of studies employing viral culture to establish the presence of replicable infectious virus,” OSF. 2023.

[18] P. F. Whiting, A. W. S. Rutjes, M. E. Westwood, and S. Mallett, “A systematic review classifies sources of bias and variation in diagnostic test accuracy studies,” J. Clin. Epidemiol., vol. 66, no. 10, pp. 1093–1104, 2013, doi: 10.1016/J.JCLINEPI.2013.05.014.

[19] Y. Akiyama et al., “A Pilot Study on Viral Load in Stool Samples of Patients with COVID-19 Suffering from Diarrhea,” Jpn. J. Infect. Dis., vol. 75, no. 1, pp. 36–40, 2022, doi: 10.7883/YOKEN.JJID.2021.018.

[20] S. Albert, A. Ruíz, J. Pemán, M. Salavert, and P. Domingo-Calap, “Lack of evidence for infectious SARS-CoV-2 in feces and sewage,” Eur. J. Clin. Microbiol. Infect. Dis., vol. 40, no. 12, pp. 2665– 2667, 2021, doi: 10.1007/S10096-021-04304-4.

[21] I. P. Ribeiro et al., “Infectious SARS-CoV-2 Particles from Rectal Swab Samples from COVID-19 Patients in Brazil,” Viruses, vol. 15, no. 5, 2023, doi: 10.3390/V15051152.

[22] R. Wölfel et al., “Virological assessment of hospitalized patients with COVID-2019,” Nature, vol. 581, no. 7809, pp. 465–469, 2020, doi: 10.1038/S41586-020-2196-X.

[23] H. Yao et al., “Patient-derived SARS-CoV-2 mutations impact viral replication dynamics and infectivity in vitro and with clinical implications in vivo,” Cell Discov., vol. 6, no. 1, 2020, doi: 10.1038/S41421-020-00226-1.

[24] C. Cerrada-Romero et al., “Excretion and viability of SARS-CoV-2 in feces and its association with the clinical outcome of COVID-19,” Sci. Rep., vol. 12, no. 1, 2022, doi: 10.1038/S41598-022-11439-7.

[25] J. Dergham, J. Delerce, M. Bedotto, B. La Scola, and V. Moal, “Isolation of Viable SARS-CoV-2 Virus from Feces of an Immunocompromised Patient Suggesting a Possible Feacal Mode of Transmission,” J. Clin. Med., vol. 10, no. 12, 2021, doi: 10.3390/JCM10122696.

[26] T. M. Fumian et al., “SARS-CoV-2 RNA detection in stool samples from acute gastroenteritis cases, Brazil,” J. Med. Virol., vol. 93, no. 4, pp. 2543–2547, 2021, doi: 10.1002/JMV.26786.

[27] H. W. Jeong et al., “Viable SARS-CoV-2 in various specimens from COVID-19 patients,” Clin. Microbiol. Infect., vol. 26, no. 11, pp. 1520–1524, 2020, doi: 10.1016/J.CMI.2020.07.020.

[28] M. Joshi et al., “Lack of evidence of viability and infectivity of SARS-CoV-2 in the feacal specimens of COVID-19 patients,” Front. public Heal., vol. 10, 2022, doi: 10.3389/FPUBH.2022.1030249.

[29] M. Lavania et al., “Prolonged Shedding of SARS-CoV-2 in Feces of COVID-19 Positive Patients: Trends in Genomic Variation in First and Second Wave,” Front. Med., vol. 9, 2022, doi: 10.3389/FMED.2022.835168.

[30] F. Nogueira et al., “Intestinal Shedding of SARS-CoV-2 in Children: No Evidence for Infectious Potential,” Microorganisms, vol. 11, no. 1, 2022, doi: 10.3390/MICROORGANISMS11010033.

[31] R. M. Pedersen et al., “Rectally shed SARS-CoV-2 in COVID-19 inpatients is consistently lower than respiratory shedding and lacks infectivity,” Clin. Microbiol. Infect., vol. 28, no. 2, pp. 304.e1–304.e3, 2022, doi: 10.1016/J.CMI.2021.10.023.

[32] Y. C. Lin et al., “Detection and quantification of infectious severe acute respiratory coronavirus-2 in diverse clinical and environmental samples,” Sci. Rep., vol. 12, no. 1, 2022, doi: 10.1038/S41598-022-09218-5.

[33] J. Zhou et al., “Viral emissions into the air and environment after SARS-CoV-2 human challenge: a phase 1, open label, first-in-human study,” The Lancet. Microbe, vol. 4, no. 8, pp. e579–e590, 2023, doi: 10.1016/S2666-5247(23)00101-5.

[34] J. Bullard et al., “Predicting Infectious Severe Acute Respiratory Syndrome Coronavirus 2 From Diagnostic Samples,” Clin. Infect. Dis., vol. 71, no. 10, pp. 2663–2666, 2020, doi: 10.1093/CID/CIAA638.

[35] N. Decaro et al., “Long-term persistence of neutralizing SARS-CoV-2 antibodies in pets,” Transbound. Emerg. Dis., vol. 69, no. 5, pp. 3073–3076, 2022, doi: 10.1111/TBED.14308.

[36] A. R. Zhang et al., “Ecology of Middle East respiratory syndrome coronavirus, 2012-2020: A machine learning modelling analysis,” Transbound. Emerg. Dis., vol. 69, no. 5, pp. e2122–e2131, 2022, doi: 10.1111/TBED.14548.

[37] Q. Qian et al., “Direct Evidence of Active SARS-CoV-2 Replication in the Intestine,” Clin. Infect. Dis., vol. 73, no. 3, pp. 361–366, 2021, doi: 10.1093/CID/CIAA925.

[38] H. Hopfer et al., “Hunting coronavirus by transmission electron microscopy - a guide to SARS-CoV-2-associated ultrastructural pathology in COVID-19 tissues,” Histopathology, vol. 78, no. 3, pp. 358–370, 2021, doi: 10.1111/HIS.14264.

[39] T. Jefferson et al., “Viral cultures, cycle threshold values and viral load estimation for assessing SARS-CoV-2 infectiousness in haematopoietic stem cell and solid organ transplant patients: a systematic review,” J. Hosp. Infect., vol. 132, pp. 62–72, 2023, doi: 10.1016/J.JHIN.2022.11.018.

[40] T. Jefferson, E. A. Spencer, J. Brassey, and C. Heneghan, “Viral Cultures for Coronavirus Disease 2019 Infectivity Assessment: A Systematic Review,” Clin. Infect. Dis., vol. 73, no. 11, pp. E3884– E3899, 2021, doi: 10.1093/CID/CIAA1764.

[41] I. J. Onakpoya et al., “SARS-CoV-2 and the role of close contact in transmission: a systematic review,” F1000Research, vol. 10, p. 280, 2022, doi: 10.12688/F1000RESEARCH.52439.3/DOI.

[42] E. A. Bruce et al., “Predicting infectivity: comparing four PCR-based assays to detect culturable SARS-CoV-2 in clinical samples,” EMBO Mol. Med., vol. 14, no. 2, 2022, doi: 10.15252/EMMM.202115290.

[43] C. A. Bryant, S. A. Wilks, and C. W. Keevil, “Survival of SARS-CoV-2 on the surfaces of food and food packaging materials,” 2022. doi: 10.46756/sci.fsa.kww583.

[44] Y. Geng et al., “Increased Incidencge and Risk Factors of Infections by Extended-Spectrum β-Lactamase-Producing Enterobacterales During the COVID-19 Pandemic: A Retrospective Case-Control Study,” Infect. Drug Resist., vol. 16, pp. 4707–4716, 2023, doi: 10.2147/IDR.S421240.

