## Appendix 1. Literature search strategy. App 2. List of excluded studies, with reasons. App 3. Additional information on the methods and results for "Oro-faecal transmission of SARS-CoV-2: A systematic review of studies employing viral culture from gastrointestinal and other potential sources"

**Literature search strategy**

We searched four main databases:

- **WHO Covid-19 Database** (<https://search.bvsalud.org/global-literature-on-novel-coronavirus-2019-ncov/>)

The global literature cited in the WHO COVID-19 database is updated daily (Monday through Friday) from searches of bibliographic databases, hand searching, and the addition of other expert-referred scientific articles. The database ceased adding manual updates in June 2023.

- **LitCovid** (<https://www.ncbi.nlm.nih.gov/research/coronavirus/>)

A curated literature hub for tracking up-to-date scientific information about the 2019 novel Coronavirus. It is a comprehensive resource on the subject, providing a central access to relevant articles in PubMed.

- **medRxiv** (<https://www.medrxiv.org/>)

A free online archive and distribution server for complete but unpublished manuscripts (preprints) in the medical, clinical, and related health sciences.

- **Google Scholar** (<https://scholar.google.com/>)

Provides a broad search for scholarly literature across many disciplines and sources: articles, theses, books, abstracts and court opinions, from academic publishers, professional societies, online repositories, universities and other web sites.

Searches were initially undertaken monthly but as the volume of new studies increased, they moved to every fortnight during the peak of the pandemic before being less sporadic. This latest update occurred Nov 28, 2023 which was a one-off update from the previous one from November 8, 2022. For databases which do not support date granularity, date of publication was approximated. All retrieved articles are entered into a google sheet that is then used to filter and check articles for inclusion.

WHO Covid-19 Database, LitCovid are specific Covid-19 databases so there was no requirement to add a search string to identify Covid-19 articles. For medRxiv and Google Scholar we used the terms *coronavirus OR covid-19 OR SARS-CoV-2*.

For the orofecal topic we used the following terms combined with the term *transmission*:

Orofecal: *orofecal OR oro-fecal OR faecal OR fecal OR stool OR faeces OR feces OR rectal OR rectum OR anal OR anus OR toilet*

Additional techniques were used to identify relevant topics:

- For articles that looked particularly relevant, citation tracking was undertaken.
- For included systematic reviews, the lists of included and excluded articles were examined for inclusion.

**Appendix 2**

**List of studies excluded at full text screening.**

Full-text articles excluded (n = 40)

Did not use viral culture on faecal samples (n = 17)

Amin N, Haque R, Rahman MZ, Rahman MZ, Mahmud ZH, Hasan R, Islam MT, Sarker P, Sarker S, Adnan SD, Akter N, Johnston D, Rahman M, Liu P, Wang Y, Shirin T, Rahman M, Bhattacharya P. Dependency of sanitation infrastructure on the discharge of faecal coliform and SARS-CoV-2 viral RNA in wastewater from COVID and non-COVID hospitals in Dhaka, Bangladesh. Sci Total Environ. 2023 Apr 1;867:161424. doi: 10.1016/j.scitotenv.2023.161424.

Arts PJ, Kelly JD, Midgley CM, Anglin K, Lu S, Abedi GR, Andino R, Bakker KM, Banman B, Boehm AB, Briggs-Hagen M, Brouwer AF, Davidson MC, Eisenberg MC, Garcia-Knight M, Knight S, Peluso MJ, Pineda-Ramirez J, Diaz Sanchez R, Saydah S, Tassetto M, Martin JN, Wigginton KR. Longitudinal and quantitative fecal shedding dynamics of SARS-CoV-2, pepper mild mottle virus, and crAssphage. mSphere. 2023 Aug 24;8(4):e0013223. doi: 10.1128/msphere.00132-23.

Decenti CE, Salvatore MA, Mancon A, Portella G, Rocca A, Vocale C, Donati S; Italian Obstetric Surveillance System COVID-19 Working Group. A large series of molecular and serological specimens to evaluate mother-to-child SARS-CoV-2 transmission: a prospective study from the Italian Obstetric Surveillance System. Int J Infect Dis. 2023 Jan;126:1-9. doi: 10.1016/j.ijid.2022.10.045

Falahi S, Bastani E, Pakzad I, Rashidi A, Abdoli A, Kenarkoohi A. Environmental Surface Contamination with SARS-CoV-2: Toilets as the Most Contaminated Surfaces in COVID-19 Referral Hospital. Hosp Top. 2023 Apr-Jun;101(2):65-72. doi: 10.1080/00185868.2021.1969870.

Fan Q, Pan Y, Wu Q, Liu S, Song X, Xie Z, Liu Y, Zhao L, Wang Z, Zhang Y, Wu Z, Guan L, Lv X. Anal swab findings in an infant with COVID-19. Pediatr Investig. 2020 Mar;4(1):48-50. doi: 10.1002/ped4.12186.

Lavani M , Shinde M, Rawal J, Kaledhonkar A, Shinde P, Chavan N, Potdar V^.^ Detection of SARS-CoV-2 RNA in stool and urine specimens of COVID-19 positive patients by Droplet Digital RT-PCR (dd RT- PCR). Abstracts of the papers presented in the international conference of Indian Virological Society, VIROCON 2022 on “Emerging and re-emerging viral infections impacting humans, animals, plants, fish and environment” held during 05–06 November, 2022 at Sher-e-Kashmir University of Agricultural Sciences and Technology of Kashmir (SKUAST-K), Srinagar (J&K), India. *VirusDis.* 34, 97–164 (2023). <https://doi.org/10.1007/s13337-023-00811-4>

Lieputra AA, Hapsari., Permatadewi CO, Kesumayadi I, Puspitasari I, Hadi P, Farhanah N, Wahyono H, Purnomo HD Clinical and Laboratory Characteristics of COVID-19 Patients with Fecal SARS-CoV-2 Detection at a Tertiary Hospital in Indonesia. Journal of Medicinal and Chemical Sciences, 2023; 6(9): 1935-1942. doi: 10.26655/JMCHEMSCI.2023.9.2

Liu D, Zhang Y, Chen D, Wang X, Huang F, Long L, Zheng X. Evaluation of the presence of SARS-CoV-2 in vaginal and anal swabs of women with omicron variants of SARS-CoV-2 infection. Front Microbiol. 2022 Nov 8;13:1035359. doi: 10.3389/fmicb.2022.1035359.

[Meghna](https://www.microbiologyresearch.org/search?value1=Nupur+Meghna&option1=author&noRedirect=true) N, [Archana](https://www.microbiologyresearch.org/search?value1=Archana+Archana&option1=author&noRedirect=true) A, [Bhushan](https://www.microbiologyresearch.org/search?value1=Divendu+Bhushan&option1=author&noRedirect=true) D, [Kumar](https://www.microbiologyresearch.org/search?value1=Abhyuday+Kumar&option1=author&noRedirect=true) A , [Asim Sarfraz](https://www.microbiologyresearch.org/search?value1=Asim+Sarfraz&option1=author&noRedirect=true) A , [Bijaya Nanda Naik](https://www.microbiologyresearch.org/search?value1=Bijaya+Nanda+Naik&option1=author&noRedirect=true) BN, [Pati](https://www.microbiologyresearch.org/search?value1=Binod+Kumar+Pati&option1=author&noRedirect=true) BK. Prevalence of SARS- CoV-2 virus in saliva, stool, and urine samples of COVID-19 patients in Bihar, India. Preprint. Access Microbiology. Aug 29 2023. [doi.org/10.1099/acmi.0.000693.v1](https://doi.org/10.1099/acmi.0.000693.v1)

Morse AG, King-Nakaoka E, Pace R, Williams JE, Meehan C, Martin M, Ley S, Barbosa-Leiker C, McGuire MA, McGuire MK. Frequency and duration of SARS-COV-2 in feces of breastfeeding mothers with and without confirmed COVID-19. 2023 Western Medical Research Conference Abstract Supplement. Journal of Investigative Medicine. 2023;71(1):NP1-NP702. doi:[10.1177/10815589221142328](https://doi.org/10.1177/10815589221142328)

Muzellina VN, Abdullah M, Kurniawan J and Rizka A. Relationship between anal swab PCR for SARS-CoV-2 with gastrointestinal clinical manifestations and severity of COVID-19 infection in Indonesia [version 2; peer review: 2 approved]. *F1000Research* 2023, **12**:358. [doi.org/10.12688/f1000research.128821.2](https://doi.org/10.12688/f1000research.128821.2)

Nagle S, Tandjaoui-Lambiotte Y, Boubaya M, Athenaïs G, Alloui C, Bloch-Queyrat C, Carbonnelle E, Brichler S, Cohen Y, Zahar JR, Delagrèverie H. Environmental SARS-CoV-2 contamination in hospital rooms of patients with acute COVID-19. J Hosp Infect. 2022 Aug;126:116-122. doi: 10.1016/j.jhin.2022.05.003.

Shinde M, Lavania M, Rawal J, Chavan N, Shinde P. Evaluation of droplet digital qRT-PCR (dd qRT-PCR) for quantification of SARS CoV-2 RNA in stool and urine specimens of COVID-19 patients. Front Med (Lausanne). 2023 Jun 26;10:1148688. doi: 10.3389/fmed.2023.1148688.

Sokolovska L, Terentjeva-Decuka A, Cistjakovs M, Nora-Krukle Z, Gravelsina S, Vilmane A, Vecvagare K, Murovska M. The presence of SARS-CoV-2 in multiple clinical specimens of a fatal case of COVID-19: a case report. J Med Case Rep. 2022 Dec 22;16(1):484. doi: 10.1186/s13256-022-03706-y

Wang J, Feng H, Zhang S, Ni Z, Ni L, Chen Y, Zhuo L, Zhong Z, Qu T. SARS-CoV-2 RNA detection of hospital isolation wards hygiene monitoring during the Coronavirus Disease 2019 outbreak in a Chinese hospital. Int J Infect Dis. 2020 May;94:103-106. doi: 10.1016/j.ijid.2020.04.024.

Wee LE, Arora S, Ko KK, Conceicao EP, Coleman KK, Tan KY, Tohid HB, Liu Q, Tung GLT, See SWJ, Suphavilai C, Ling ML, Venkatachalam I. Environmental contamination and evaluation of healthcare-associated SARS-CoV-2 transmission risk in temporary isolation wards during the COVID-19 pandemic. Am J Infect Control. 2023 Apr;51(4):413-419. doi: 10.1016/j.ajic.2022.09.004

Zhang Z, Li X, Lyu K, Zhao X, Zhang F, Liu D, Zhao Y, Gao F, Hu J, Xu D. Exploring the Transmission Path, Influencing Factors and Risk of Aerosol Transmission of SARS-CoV-2 at Xi'an Xianyang International Airport. Int J Environ Res Public Health. 2023 Jan 3;20(1):865. doi: 10.3390/ijerph20010865.

Did not assess oroFaecal transmission: (n = 7)

Berengua C, López M, Esteban M, Marín P, Ramos P, Cuerpo MD, Gich I, Navarro F, Miró E, Rabella N. Viral culture and immunofluorescence for the detection of SARS-CoV-2 infectivity in RT-PCR positive respiratory samples. J Clin Virol. 2022 Jul;152:105167. doi: 10.1016/j.jcv.2022.105167. Epub 2022 Apr 28. PMID: 35523105; PMCID: PMC9046102.

Kojima N, Mores C, Farsai N, Klausner J. Duration of SARS-CoV-2 viral culture positivity among different specimen types. Clin Microbiol Infect. 2021 Oct;27(10):1540-1541. doi: 10.1016/j.cmi.2021.07.004. Epub 2021 Jul 16. PMID: 34274524; PMCID: PMC8282432.

Li H, Ren L, Zhang L, Wang Y, Guo L, Wang C, Xiao Y, Wang Y, Rao J, Wang X, Liu Y, Huang C, Gu X, Fan G, Li H, Lu B, Cao B, Wang J. High anal swab viral load predisposes adverse clinical outcomes in severe COVID-19 patients. Emerg Microbes Infect. 2020 Dec;9(1):2707-2714. doi: 10.1080/22221751.2020.1858700.

Rajindra Napit, Prajwol Manandhar, Ashok Chaudhary, Bishwo Shrestha, Ajit Poudel, Roji Raut, Saman Pradhan, Samita Raut, Sujala Mathema, Rajesh Rajbhandari, Sameer Dixit, Jessica S. Schwind, Christine K Johnson, Jonna K Mazet, Dibesh Karmacharya. Rapid genomic surveillance of SARS-CoV-2 in a dense urban community using environmental (sewage) samples.medRxiv 2021.03.29.21254053; doi: <https://doi.org/10.1101/2021.03.29.21254053>

Sharma DK, Nalavade UP, Kalgutkar K, Gupta N, Deshpande JM. SARS-CoV-2 detection in sewage samples: Standardization of method & preliminary observations. Indian J Med Res. 2021 Jan & Feb;153(1 & 2):159-165. doi: 10.4103/ijmr.IJMR_3541_20

Siedner MJ, Boucau J, Gilbert RF, Uddin R, Luu J, Haneuse S, Vyas T, Reynolds Z, Iyer S, Chamberlin GC, Goldstein RH, North CM, Sacks CA, Regan J, Flynn JP, Choudhary MC, Vyas JM, Barczak AK, Lemieux JE, Li JZ. Duration of viral shedding and culture positivity with postvaccination SARS-CoV-2 delta variant infections. JCI Insight. 2022 Jan 25;7(2):e155483. doi: 10.1172/jci.insight.155483. PMID: 34871181; PMCID: PMC8855795.

Zhang D, Zhang X, Yang Y, Huang X, Jiang J, Li M, Ling H, Li J, Liu Y, Li G, Li W, Yi C, Zhang T, Jiang Y, Xiong Y, He Z, Wang X, Deng S, Zhao P, Qu J. SARS-CoV-2 spillover into hospital outdoor environments. J Hazard Mater Lett. 2021 Nov;2:100027. doi: 10.1016/j.hazl.2021.100027.

Modelling study: (n = 2)

Acosta N 2021	Wastewater Monitoring of SARS-CoV-2 from Acute Care Hospitals Identifies Nosocomial Transmission and Outbreaks	<https://www.medrxiv.org/content/10.1101/2021.02.20.21251520v1>

Joob B, Wiwanitkit V. Fecal-oral transmission of COVID-19. Pediatr Investig. 2020 Jun 24;4(2):152. doi: 10.1002/ped4.12203. PMID: 32835178; PMCID: PMC7331445.

Case report: (n = 2)

Zhang Y, Chen C, Zhu S, Shu C, Wang D, Song J, Song Y, Zhen W, Feng Z, Wu G, Xu J, Xu W. Isolation of 2019-nCoV from a stool specimen of a laboratory-confirmed case of the Coronavirus Disease 2019 (COVID-19). China CDC Wkly. 2020 Feb 21;2(8):123-124.

Zhou P, Yang XL, Wang XG. et al. A pneumonia outbreak associated with a new coronavirus of probable bat origin. Nature 579, 270–273 (2020). https://doi.org/10.1038/s41586-020-2012-7.

Review: (n = 10)

Abd EW, Eassa SM, Metwally M, Al-Hraishawi H, Omar SR. SARS-CoV-2 Transmission Channels: A Review of the Literature. MEDICC Rev. 2020 Oct;22(4):51-69. doi: 10.37757/MR2020.V22.N4.3.

Jones DL, Baluja MQ, Graham DW, Corbishley A, McDonald JE, Malham SK, Hillary LS, Connor TR, Gaze WH, Moura IB, Wilcox MH, Farkas K. Shedding of SARS-CoV-2 in feces and urine and its potential role in person-to-person transmission and the environment-based spread of COVID-19. Sci Total Environ. 2020 Dec 20;749:141364. doi: 10.1016/j.scitotenv.2020.141364.

Lowry SA, Wolfe MK, Boehm AB . Respiratory virus concentrations in human excretions that contribute to wastewater: A systematic review. medRxiv 2023.02.19.23286146; doi: https://doi.org/10.1101/2023.02.19.23286146

Meyerowitz EA, Richterman A, Gandhi RT, Sax PE. Transmission of SARS-CoV-2: A Review of Viral, Host, and Environmental Factors. Ann Intern Med. 2021 Jan;174(1):69-79. doi: 10.7326/M20-5008.

Ning T, Liu S, Xu J, Yang Y, Zhang N, Xie S, Min L, Zhang S, Zhu S, Wang Y. Potential intestinal infection and faecal-oral transmission of human coronaviruses. Rev Med Virol. 2022 May 18:e2363. doi: 10.1002/rmv.2363.

Parida VK, Saidulu D, Bhatnagar A, Gupta AK, Afzal MS. A critical assessment of SARS-CoV-2 in aqueous environment: Existence, detection, survival, wastewater-based surveillance, inactivation methods, and effective management of COVID-19. Chemosphere. 2023 Jun;327:138503. doi: 10.1016/j.chemosphere.2023.138503.

Rhee C, Kanjilal S, Baker M, Klompas M. Duration of Severe Acute Respiratory Syndrome Coronavirus 2 (SARS-CoV-2) Infectivity: When Is It Safe to Discontinue Isolation? Clin Infect Dis. 2021 Apr 26;72(8):1467-1474. doi: 10.1093/cid/ciaa1249.

SAGE 2020. SARS-CoV-2 transmission routes and environments. SAGE, 22 October 2020 <https://assets.publishing.service.gov.uk/government/uploads/system/uploads/attachment_data/file/933225/S0824_SARS-CoV-2_Transmission_routes_and_environments.pdf>

Sehmi P, Cheruiyot I. Presence of Live SARS-CoV-2 Virus in Feces of Coronavirus Disease 2019 (COVID-19) Patients: A Rapid Review. MedRxiv 2020. Doi: <https://doi.org/10.1101/2020.06.27.2010542>

Termansen MB, Frische S. Fecal-oral transmission of SARS-CoV-2: A systematic review of evidence from epidemiological and experimental studies. Am J Infect Control. 2023 Dec;51(12):1430-1437. doi: 10.1016/j.ajic.2023.04.170.

Did not verify replication-competent virus in culture ( n=1)

Wang W, Xu Y, Gao R, et al. Detection of SARS-CoV-2 in different types of clinical specimens. JAMA. 2020;323(18):1843–1844. doi:10.1001/jama.2020.3786

Abstract Thesis (n=1)

Donovan J. SARS-CoV-2 infectivity potential in municipal wastewater: Implications for public health & water treatment. MSc Thesis. ["SARS-CoV-2 infectivity potential in municipal wastewater: Implications" by Justin Donovan (uwo.ca)](https://ir.lib.uwo.ca/etd/9577/).

**Appendix 3**

Methods

We conducted searches in the WHO Covid-19 Database, LitCovid, medRxiv, and Google Scholar for SARS-CoV-2 using keywords and associated synonyms, with a search date up to 8th November 2023. The searches were conducted by an information specialist (JB); no language restrictions were imposed. Forward citation screening was done to identify any additional relevant studies (see Appendix 1. Literature search strategy). Hand searches of the bibliographies of relevant studies were performed. Two reviewers (SG, ES) independently screened study abstracts to determine eligibility; disagreements were resolved through discussion, and a third reviewer (CH) was available to arbitrate if needed.

Data extraction

Using a customised data extraction sheet, we extracted the following information from included studies, where available: study design, calendar data/period when the study was done, study characteristics, setting, country, population studied, patient numbers, sample numbers, patient characteristics, clinical information, data on symptoms of the study participants including timing of symptom assessment, symptoms reported, signs observed, methods used for sample collection, numbers of samples taken, the frequency and timing of samples, methods and results of sample analysis including RT-PCR (use of internal controls, the platform with gene targets used, cycle thresholds), genome sequencing, and viral culture methods (timing, media, verification techniques, quantification). One reviewer (SG) extracted data from the included studies, with independent verification by a second reviewer (ES or JC).

Risk of bias assessment

We assessed the risk of bias within five domains, modified and extended from the QUADAS-2 criteria [1]. The risk of bias domains was assessed as follows: 1) were the criteria for diagnosing a case clearly reported and appropriate?; 2) was the reporting of patient/ population characteristics adequate?; 3) were the methods used to obtain RT-PCR results replicable and appropriate?; 4) were the methods used to obtain viral culture results replicable and appropriate?; and 5) were the analysis and reporting of the results appropriate?

Three reviewers (SG, ES, JC) independently assessed the risk of bias, and another independent reviewer (CH) was available to arbitrate in case of disagreement.

Data reporting and analysis

We followed PRISMA reporting guidelines for systematic reviews [2]. From the included studies, we extracted data as detailed above. We generated tables to present the main results of the included studies plus the risk of bias and included a forest plot as appropriate to show the proportions of positive samples with 95% confidence intervals from the included studies.

Descriptive Assessment of the Individual Studies for Risk of Bias

Akiyama 2022 [3] reported data from 13 patients, with an age range from 21 to 83 years, three females and 10 males. Two patients had severe disease and three had diarrhea. Disease severity was critical in 3 patients, severe in 4 patients, moderate in 6 patients. Of the eight samples tested with cell culture in Vero E6/TMPRESS2 cells over 4 weeks, no SARS-CoV-2 replication was observed. The authors also described results from four COVID-19 patients with SAS-Cov-2 RNA positive feacal samples. The first patient had a positive stool sample on day 18 with a viral load of 3.51 log10 copies/g, that increased at day 22 to 7.83 log10 copies/g, at day 24 to 8.28 log10 copies/g, and became negative on day 26. The second patient presented a positive stool on day 16 with a viral load of 4.16 log10 copies/g and a negative sample on day 34. The third patient had a positive stool sample at day 20 with a viral load of 5.36 log10 copies/g, that decreased to 3.20 log10 copies/g at day 30. The fourth patient had a positive stool sample at day 2 with a 5.17 log10 copies/g, that decreased to 4.39 log10 copies/g at day six. Despite the high genome copy numbers, none of the eight samples was found to be positive by cell culture using Vero E6/TMPRESS2 cells over four weeks.

No information was reported regarding how the selection of samples subjected to viral culture was carried out and the methods used to identify viruses growing in cell culture.

Albert et al. [4] reported that 26 feacal samples from 8 patients were available, each apparently tested by RT-PCR in duplicate and we have reported the mean value for each paired result. Among the 26 samples tested by RT-PCR, SARS-CoV-2 was detected in 16 (61.5%). Although the Ct cut-off used to indicate “undetectable” was not specified in the paper, among the 16 with positive results, Cts ranged from 34.2 to 42.7, indicating that the Ct cut-off point used must have been higher than 42.7 (suggesting very low concentrations of SARS-CoV-2 RNA or background noise from contaminating materials in the extraction process). It was not clear how samples were selected for cell culture; the report states “Feacal and sewage samples with the highest RNA detected by RT-qPCR were used to inoculate Vero E6 cells…”. Eight feacal samples from six patients were tested by cell culture, and none showed evidence of viral replication. They report using RT-qPCR on the cell culture supernatant to assess if there was an increase over time, which would have indicated SARS-CoV-2 replication in culture. Among the 8 patients, 5 were reported to have respiratory symptoms, and 3 had gastrointestinal symptoms; there was no data on time from symptom onset or how that related to the schedule of sample collection. The study did not report results from testing respiratory samples.

Cerrada-Romero and colleagues [5] reported 27 of the 62 patients (from whom 79 samples were collected) tested positive for SARS-CoV-2 RNA in their feaces using RT-PCR. The Ct values reported were a median 31.2 (range 24.5 to 39.6) for samples collected within the first week of hospital admission, and median 34.5 (27.2 to 39.1) for samples collected in the second week of hospital admission. A total of 31 RNA-positive samples were selected for testing by viral cell culture; details of how these were chosen were not reported. Undiluted stool samples and serial dilutions up to 1:16 were added to 96-well microplates containing Vero E6 cell monolayers. Incubation was without CO2. The supernatant was tested for the presence of SARS-CoV-2 RNA using PCR: none of the cell culture supernatants were positive for SARS-CoV-2. Pneumonia was detected on admission in all COVID-19 patients whose stool samples tested negative by PCR, and 75% of those whose stool samples were tested were positive by PCR. Within contemporaneous nasopharyngeal swabs, there was no difference in viral load between patients with positive or negative stool samples. Nasopharyngeal swabs were negative in 3/9 and 3/6 patients tested in weeks 3 to 4 and 2 to 4 months, respectively, after symptom onset.

Dergham et al. [6] reported that 128 stool samples obtained from 59 patients tested positive by RT-PCR, and from these, 106 samples from 46 patients were available for cell culture. After three rounds of cell culture, two of the samples, both obtained from an immunocompromised patient, displayed cytopathic effect (CPE). Confirmatory identification with qPCR for SARS-CoV-2 was done followed by whole genome sequencing (WGS) on the cell culture supernatant. Genome sequencing of the cell supernatants of the two CPE-positive cultures indicated that two separate strains were present in the two samples from the same immunocompromised patient. This patient had diabetes, hypertension and obesity and had had a kidney transplant 21 years earlier. On admission, pneumonia was diagnosed by chest computed tomography; however, a positive RT-PCR from respiratory swabs only occurred two days later and with a high Ct value. Dergham et al. reported observing a cytopathic effect after three rounds of culture of two feacal samples (with confirmation of SARS-CoV-2 RNA in the culture supernatant) and it is not clear why three rounds of culture were done, as SARS-CoV-2 in culture forms clear, easily visible plaques and it is typical for plaques to be evident on the first plating [7]. This observation could potentially indicate that there was a very low quantitative presence of virus in culture or the virus was present but not capable of growth until it was subcultured due to large amounts of incompetent virus and it required the dilution of inhibitors to allow for competent growth on cell culture. However, for the two samples for which viral culture was successful and for which cycle counts were reported, the RT-PCR cycle counts were 17.29 and 16.22, which supports the actual presence of infectious virus within the feacal samples from this immunocompromised patient. Immunocompromised individuals are not typical in their clinical course of SARS-CoV-2 infection, often showing prolonged viral shedding and cyclical resurgence and subsidence of symptoms over a period of months. This finding would be in line with other published findings on immunocompromised individuals [8].

The included study that reported visual evidence of SARS-CoV-2 using electron microscopy provides uncertain evidence of the presence of replication-competent SARS-CoV-2 [9]. Identifying SARS-CoV-2 through electron microscopy (the only method available to visualise assembled SARS-CoV-2 virions within tissues) is hampered by subcellular structural changes mimicking coronaviruses, by disruptions being vulnerable to damage during tissue preservation, and by the timing of tissue collection being critical; previous reports of virions visualised in tissues outside the respiratory tract and lungs have been contested[10].

Fumian et al reported that among 29 samples, 8 (27.5%) were positive when tested by RT-PCR [11]. Ct values were reported as: N1 primer: mean 34.1 (range 29.8 to 37.0); N2 primer: 5 not detected; among 5 positive cases, mean 35.5 (range 34.3 to 37.8). Four RNA-positive samples with low Ct values (from 29.8 to 32.4) were submitted to virus isolation in Vero E6 cell cultures. In cell culture, none of the four showed any evidence of CPE. CPE was evaluated using a microscope, and cell supernatant from the three passages was obtained, the RNA extracted and tested by RT-PCR for the presence of SARS-CoV-2, but none was positive for SARS-CoV-2 RNA. All included patients had acute gastroenteritis. This study used a cohort defined by the presence of gastrointestinal symptoms, and no data were available on respiratory samples or the course of the disease.

Jeong et al. [12] reported log copy numbers for feacal samples from three patients: patient 2: 1.17 +/- 0.32 log copies; patient 4: 2.18 +/- 0.11 log copies; patient 5: 2.01 +/- 0.28 log copies. Criteria for the selection of samples for viral culture was not reported. Cells were monitored daily for CPE. RT-PCR was performed on cell supernatants of infected cultures. Of three samples tested, none showed evidence of positive cell culture. The authors report that the clinical symptoms had resolved completely at the time of sampling for all patients except one. One patient was on day 15 of illness and was on a ventilator and extracorporeal membrane oxygenation (ECMO) support. Among the four clinically recovered patients from whom feacal samples were taken, viral load in the feacal samples was equal to or higher than in nasopharyngeal samples taken at the same time; in the samples taken from the critically ill patient, day 15 into the illness, viral load was higher in the feacal sample than in the corresponding nasopharyngeal swab.

Joshi [13] presented a multicenter case-series from hospitals in India using stored samples collected between May 2020 and August 2021. Specimens were collected between 0 and 6 days post onset and the viral RNA load in the feacal specimens was estimated using N gene based digital RT-PCR assay. Analysis of the viral RNA load in feacal specimens indicated 5526.2–15.2 copies per/μL of RNA. Feacal specimens of COVID-19 patients were subjected to RT-PCR, digital RT-PCR, Next Generation Sequencing (NGS), isolation in Vero CCL-81 cell lines. Ct values of the feacal specimens ranged from 19.5–25.9 and 22.6–28.1 for E gene and ORF region of SARS-CoV-2, respectively. Among the ten patients enrolled, six were experiencing symptoms of fever, cough, sore throat, chest pain, wheezing, abdominal pain, vomiting/nausea, diarrheoa, loss of taste and smell, while the remaining four were asymptomatic. None of them was found positive for replicating virus on culture. All feacal specimens had viral loads < 10^4^ copies /ml which is below the usual threshold to detect culturable virus.

Lavania and colleagues [14] collected feacal samples obtained from patients with confirmed COVID-19 admitted to hospital with symptoms of myalgia, sore throat, fever, and cough hypoxia; 15.4% showed GI symptoms, including diarrheoa and abdominal pain. 62 (67.39%) patients, who tested positive for SARS-CoV2 RNA in feaces, were identified as asymptomatic cases. Overall in this study, among feacal samples tested by RT-PCR, 173/280 (61.8%) patients tested positive. 108 (38.5%) patients were mild cases: 60/108 of whom tested positive for viral RNA in feaces; 48/108 tested negative. 80 patients were severe cases with hypoxia, SPO2 <94%. 51/80 tested positive for SARS-CoV-2 viral RNA in feaces; 29/80 tested negative. There was no association between GI symptoms, severity, and feacal viral load. The median duration between symptom onset and the first positive RT-PCR feacal test result was 11 (IQR 7 to 13) days. Respiratory sample data for these patients were not reported. The authors reported that “Ct values in the feacal specimens were lower than in the throat specimens”. Fifty-five samples with “low Ct” (cycle count numbers not reported) were selected for viral culture attempts. Cell culture was inspected daily for CPE, and cell supernatants were taken from passage 1 on day 2 and from passage 2 on day 4 and tested using RT-PCR for SARS-CoV-2. The authors report that virus isolation was not successful from any sample; also that they additionally tested feacal samples for potential replicability of SARS-CoV-2 by amplifying subgenomic RNA, but the methods and results from these subgenomic assays are incompletely reported.

Nogueira et al [15] presented data from three clinical centers in Austria regarding 206 hospitalised children diagnosed between December 2020 and June 2021. Aliquots of positive samples were kept at 4^0^ C (which preserves virus infectivity better than dissolving in 0.9% NaCl and/or freezing at -80^0^ C) for further testing of viral viability in cell culture. The Vero-E6 (ATCC CRL-1586) cell line, which is permissive for the propagation of SARS-CoV-2, was used in the present study. The SARS-CoV-2 BU2-NS P3 strain was used as a positive control, while non-infected Vero-E6 cells (mock) served as a negative control. The study included mainly patients for elective surgery, with gastrointestinal symptoms, and various other problems mostly unrelated to acute infection. None of the seven feacal samples analysed was found positive on culture.

Pedersen et al. [16] reported data from 28 COVID-19 patients with 33 pairs of oropharyngeal and rectal swabs analysed. Four patients provided more than one pair of samples. A total of 11 patients had diarrheoa, 6 patients had nausea/ vomiting and 8 patients had received proton pump inhibitors at the time of admission. Of 33 rectal swabs, 14 were found to be positive, with a median concentration of SARS-CoV-2 RNA of 13,014 IU/ml (IQR 6824 to 34,403). Supernatants analysed by RT-PCR on inoculation and on days 3 and 6 to detect viral proliferation and confirm viral identity found that none of the 33 samples contained culturable virus. In the corresponding respiratory samples, 11/33 samples contained culturable virus. The paired rectal sample was negative by RT-PCR for three of the culturable respiratory samples.

Ribeiro et al [17] presented results from 333 rectal swabs from 130 persons from a slum in Rio de Janeiro, Brazil, from May to October 2020. Some of them were symptomatic and others were their household contacts. Rectal samples provided evidence of active SARS-CoV-2 replication in intestinal cells. As sg mRNA is transcribed only in infected cells and is not packaged into virions, its detection indicates the presence of actively infected cells in samples. The rectal swab was collected from a 68-year-old woman who tested positive for SARS CoV-2, with a Ct value of 25.2. A cytopathic effect (CPE) was observed after 3 days of incubation, and the Ct value of the infected-culture supernatant was 9.1. A second-round passage, performed three days post-infection, generated a viral stock. Presence of infectious SARS-CoV-2 was found from 1 out of 14 rectal swab samples inoculated in Vero cells. They used an excellent technique but rectal swabs were expected to have less levels of contamination. 43/333 (12.9%) rectal swabs were RT PCR + representing 33 patients. Ct-values of positive-RS samples ranged from 25.2 to 36.4, with a median value of 33.5. Subgenomic N mRNA was detected in 6/31 (19.1%) rectal swab samples.

Wölfel et al. [18] reported on a series of 9 mild COVID-19 cases, including analysis of respiratory and non-respiratory samples. RT-PCR of cell culture supernatants at a series of time points was used to investigate the presence of replicating SARS-CoV-2. They did not report the results (Ct values or log copies) of RT-PCR tests of feacal samples, nor how samples were selected for testing by viral culture; they reported that no infectious virus was isolated from 13 stool samples obtained from 4 patients: They reported successfully isolating SARS-CoV-2 in several nasopharyngeal and sputum samples obtained from the same patients less than 8 days after symptoms began.

Yao et al analysed [19] stool samples from 11 patients and found three positive stool samples from severe disease patients treated in hospital (not in intensive care), who later recovered. The first patients had their symptom onset at the 10th day, the second at 15th day and the third at 16th day. Respiratory sample data were not available for the patients from whom stool samples were subjected to viral culture. They reported finding viable viral isolates in cell culture from three stool samples, with Cts ≤ 28 from the culture supernatant and a decrease in Ct over time shown by RT-PCR analysis. It is unclear how the samples were chosen and what corresponding respiratory samples showed.

References

[1]	P. F. Whiting, A. W. S. Rutjes, M. E. Westwood, and S. Mallett, “A systematic review classifies sources of bias and variation in diagnostic test accuracy studies,” *J Clin Epidemiol*, vol. 66, no. 10, pp. 1093–1104, 2013, doi: 10.1016/J.JCLINEPI.2013.05.014.

[2]	M. J. Page *et al.*, “The PRISMA 2020 statement: an updated guideline for reporting systematic reviews,” *BMJ*, vol. 372, Mar. 2021, doi: 10.1136/BMJ.N71.

[3]	Y. Akiyama *et al.*, “A Pilot Study on Viral Load in Stool Samples of Patients with COVID-19 Suffering from Diarrhea,” *Jpn J Infect Dis*, vol. 75, no. 1, pp. 36–40, 2022, doi: 10.7883/YOKEN.JJID.2021.018.

[4]	S. Albert, A. Ruíz, J. Pemán, M. Salavert, and P. Domingo-Calap, “Lack of evidence for infectious SARS-CoV-2 in fecal and sewage,” *Eur J Clin Microbiol Infect Dis*, vol. 40, no. 12, pp. 2665–2667, Dec. 2021, doi: 10.1007/s10096-021-04304-4 [doi].

[5]	C. Cerrada-Romero *et al.*, “Excretion and viability of SARS-CoV-2 in feces and its association with the clinical outcome of COVID-19,” *Sci Rep*, vol. 12, no. 1, Dec. 2022, doi: 10.1038/S41598-022-11439-7.

[6]	J. Dergham, J. Delerce, M. Bedotto, B. La Scola, and V. Moal, “Isolation of Viable SARS-CoV-2 Virus from Feces of an Immunocompromised Patient Suggesting a Possible Fecal Mode of Transmission,” *J Clin Med*, vol. 10, no. 12, Jun. 2021, doi: 10.3390/JCM10122696.

[7]	Y. C. Lin *et al.*, “Detection and quantification of infectious severe acute respiratory coronavirus-2 in diverse clinical and environmental samples,” *Sci Rep*, vol. 12, no. 1, Dec. 2022, doi: 10.1038/S41598-022-09218-5.

[8]	I. Rajakumar *et al.*, “Extensive environmental contamination and prolonged severe acute respiratory coronavirus-2 (SARS CoV-2) viability in immunosuppressed recent heart transplant recipients with clinical and virologic benefit with remdesivir,” *Infect Control Hosp Epidemiol*, vol. 43, no. 6, pp. 817–819, Jun. 2022, doi: 10.1017/ICE.2021.89.

[9]	C. Zhang *et al.*, “Clinical and epidemiological characteristics of pediatric SARS-CoV-2 infections in China: A multicenter case series,” *PLoS Med*, vol. 17, no. 6, Jun. 2020, doi: 10.1371/JOURNAL.PMED.1003130.

[10]	H. Hopfer *et al.*, “Hunting coronavirus by transmission electron microscopy - a guide to SARS-CoV-2-associated ultrastructural pathology in COVID-19 tissues,” *Histopathology*, vol. 78, no. 3, pp. 358–370, Feb. 2021, doi: 10.1111/HIS.14264.

[11]	T. M. Fumian *et al.*, “SARS-CoV-2 RNA detection in stool samples from acute gastroenteritis cases, Brazil,” *J Med Virol*, vol. 93, no. 4, pp. 2543–2547, Apr. 2021, doi: 10.1002/JMV.26786.

[12]	H. W. Jeong *et al.*, “Viable SARS-CoV-2 in various specimens from COVID-19 patients,” *Clin Microbiol Infect*, vol. 26, no. 11, pp. 1520–1524, Nov. 2020, doi: 10.1016/J.CMI.2020.07.020.

[13]	M. Joshi *et al.*, “Lack of evidence of viability and infectivity of SARS-CoV-2 in the fecal specimens of COVID-19 patients,” *Front Public Health*, vol. 10, Oct. 2022, doi: 10.3389/FPUBH.2022.1030249.

[14]	M. Lavania *et al.*, “Prolonged Shedding of SARS-CoV-2 in Feces of COVID-19 Positive Patients: Trends in Genomic Variation in First and Second Wave,” *Front Med (Lausanne)*, vol. 9, Mar. 2022, doi: 10.3389/FMED.2022.835168.

[15]	F. Nogueira *et al.*, “Intestinal Shedding of SARS-CoV-2 in Children: No Evidence for Infectious Potential,” *Microorganisms*, vol. 11, no. 1, Jan. 2022, doi: 10.3390/MICROORGANISMS11010033.

[16]	R. M. Pedersen *et al.*, “Rectally shed SARS-CoV-2 in COVID-19 inpatients is consistently lower than respiratory shedding and lacks infectivity,” *Clin Microbiol Infect*, vol. 28, no. 2, pp. 304.e1-304.e3, Feb. 2022, doi: 10.1016/J.CMI.2021.10.023.

[17]	I. P. Ribeiro *et al.*, “Infectious SARS-CoV-2 Particles from Rectal Swab Samples from COVID-19 Patients in Brazil,” *Viruses*, vol. 15, no. 5, May 2023, doi: 10.3390/V15051152.

[18]	R. Wölfel *et al.*, “Virological assessment of hospitalized patients with COVID-2019,” *Nature*, vol. 581, no. 7809, pp. 465–469, May 2020, doi: 10.1038/S41586-020-2196-X.

[19]	H. Yao *et al.*, “Patient-derived SARS-CoV-2 mutations impact viral replication dynamics and infectivity in vitro and with clinical implications in vivo,” *Cell Discov*, vol. 6, no. 1, Dec. 2020, doi: 10.1038/S41421-020-00226-1.
