## Supplementary material for "Oro-faecal transmission of SARS-CoV-2: A systematic review of studies employing viral culture from gastrointestinal and other potential sources": Table S2

Table S2. Risk of bias assessment results.

|  | Were the criteria for diagnosing a case clearly reported and appropriate? | Was the reporting of patient/ population characteristics adequate? | Were the methods used to obtain RT-PCR results replicable and appropriate? | Was the study period, including follow-up, sufficient to investigate orofecal transmission? | Were the methods used to obtain viral culture results replicable and appropriate? | Were the analysis and reporting of the results appropriate |
| --- | --- | --- | --- | --- | --- | --- |
| Akiyama, 2022 | yes | yes | yes | N/A | unclear | yes |
| Albert 2021 | yes | yes | unclear | N/A | unclear | yes |
| Cerrada-Romero C 2022 | yes | yes | yes | yes | unclear | yes |
| Dergham J 2021 | yes | yes | yes | yes | unclear | yes |
| Fumian TM 2021 | yes | yes | yes | yes | unclear | yes |
| Jeong 2020 | yes | yes | unclear | N/A | unclear | unclear |
| Joshi 2022 | yes | yes | yes | N/A | yes | yes |
| Lavania M 2022 | yes | yes | yes | yes | unclear | yes |
| Nogueira 2022 | yes | yes | yes | N/A | yes | yes |
| Pedersen 2022 | yes | yes | yes | N/A | yes | yes |
| Ribeiro 2022 | yes | yes | No | N/A | yes | yes |
| Wang W 2020 | yes | unclear | yes | N/A | unclear | yes |
| Wölfel 2020 | yes | no | unclear | N/A | unclear | yes |
| Yao 2020 | yes | yes | yes | N/A | yes | unclear |
